# Multi-Site Analysis of Biosynthetic Gene Clusters (BGCs) from the Periodontitis Oral Microbiome

**DOI:** 10.1101/2023.03.02.23286703

**Authors:** Mohamad Koohi-Moghadam, Rory M. Watt, W. Keung Leung

## Abstract

Bacteria are key modulators of human health and disease. Biochemicals synthesized by bacterial biosynthetic gene clusters (BGCs) have been shown to play key roles in microbiome–host and microbe–microbe interactions. Whilst BGCs have been widely investigated in the human gut, very few studies have explored BGCs within oral niches. In this pilot study, we used shotgun metagenomic sequencing to profile the microbiota from three different oral sites: saliva, subgingival plaque, and supragingival plaque, within subjects with periodontitis (n = 23) versus controls (n = 16). Our aim was to identify BGCs associated with periodontitis, as well as BGCs that exhibited site (niche) selectivity. We identified 10,742 BGCs in the cohort, some of which were unique for a specific oral site. Aryl polyenes (APEs) and Bacteriocin were the most prevalent clusters, but we also found several ‘novel’ BGCs that were widely distributed across diverse bacterial phyla; other BGCs exhibited selectivity for periodontitis niches. Taken together, our findings significantly expand our metagenomic understanding of microbiota within healthy and diseased oral niches. By uncovering previously unexplored biosynthetic pathways, we provide a potential road-map for the future exploration of poorly understood host-microbe and microbe-microbe interactions in the oral cavity that may contribute to periodontitis.

## Introduction

Bacteria are the dominant form of life that can inhabit different human body sites such as the oral cavity, gut, upper respiratory tract, urogenital tract and skin. These ‘simple’ organisms play important roles in a myriad of physiological processes, i.e., immune responses, metabolic pathways, and nutrient cycling [1]. Studies show bacteria living on/in human hosts can either predispose someone to disease or promote health [2]. For example, shifts in microbiome composition can potentially induce diseases such as periodontal disease, dental caries, obesity, inflammatory bowel disease, diabetes, and colorectal cancer [3, 4]. On the other hand, certain bacterial taxa such as *Bifidobacterium animalis* may boost the human immune system [5].

The exotic roles of bacteria can be potentially associated with the presence of bacterial biosynthetic gene clusters (BGCs). BGCs are groups of genes located together on the bacterial genome, whose encoded proteins functionally-cooperate to produce biochemicals known as secondary metabolites [6]. Many of these secondary metabolites interact with host proteins to influence the host’s biological pathways. In addition, bacteria use these small molecules to communicate directly with each other or their wider environment [7]. Thus, bacterial BGCs play important roles in both host-microbe and microbe-microbe interactions. Thanks to advancing technologies spanning next-generation sequencing (NGS), high throughput mass spectrometry, and efficient computational biology algorithms, new BGCs and their associated secondary metabolites can be identified in a time- and cost-efficient manner. For example, almost 1,000 putative BGCs were discovered in 169 *Streptococcus mutans* genomes, demonstrating an amazing capacity to generate small compounds within a single bacterial species [8, 9].

The human oral cavity comprises several distinct bacterial habitats, including teeth (both above and below the ‘gum-line’), oral implants, oral appliances (e.g. dentures), gingivae (gums), tongue dorsum, palatine tonsils, throat, and saliva; which may each host site-specific bacterial taxa [10]. The oral cavity is exposed to the outside environment and can be considered one of the most accessible sites for putative pathogens or commensals. Exogenous colonizers can potentially shift the healthy homeostatic balance (eubiosis) of oral bacterial communities. A dysbiosis in oral bacterial communities may potentially induce diseases such as caries or periodontitis [11].

Here, we used a systematic approach to analyze oral metagenome samples to survey oral BGCs. We collected samples from three different oral sites, namely, saliva, subgingival plaque and supragingival plaque, from 39 dentated Chinese individuals (23 subjects with periodontitis and 16 controls). From these samples, we discovered over 10,000 BGCs, some of which are unique to a particular oral site and associated with periodontal diseases. Our findings help to build a comprehensive profile of BGCs associated with periodontitis in the oral cavity of a Chinese cohort.

## Materials and method

### Ethics statement

This study was approved by the Institutional Review Board (IRB) of the University of Hong Kong/Hospital Authority Hong Kong West Cluster (approval number HKU/UW 20-364). All subjects were given verbal explanation regarding the investigation upon first contact. Written informed consent was obtained from all participants who agreed to take part. Personal identifiers were removed from all collected data. All procedures performed were in accordance with the ethical standards of the institutional research committee and with the Declaration of Helsinki revised in 2013.

### Subject recruitment

Inclusion criteria were: i) adults ≥ 18 years old; ii) in good general health and did not have any medical conditions that could potentially affect their oral microbiome; iii) without self-reported salivary gland disease or dry mouth of any type. Subjects were excluded if they were: i) smokers, ii) on medication, ii) had systemic disease, iii) pregnant, or iv) undergoing orthodontic treatment.

### Clinical Examination

Recruitment, consent and clinical examination were carried out at the Reception and Primary Care Clinic, Faculty of Dentistry, the University of Hong Kong between 1/10/2021-30/4/2022. All examinations were carried out by two calibrated examiners under the direction of WKL. Examinations were repeated in every tenth participant to assess inter-examiners’ reliability. Participants provided written consent underwent a clinical examination, which was carried out by two calibrated examiners. Due to the coronavirus disease (COVID) pandemic, all participants were prescribed a hydrogen peroxide based pre-procedural mouth rinse on every attendance for cross-infection risk-mitigation [12]. As described previously [13], DMFT index [14] was used to evaluate dental conditions, with. decayed teeth defined at International Caries Detection and Assessment System (ICDAS) code 3 [localized enamel breakdown (without clinical visual signs of dentinal involvement)] or above [15, 16].

We used a periodontal probe (PCP-UNC 15, HuFriedy Manufacturing Co., Chicago, IL, USA) to record periodontal parameters from six sites of each tooth (mesio-buccal, mid-buccal, disto-buccal, mesio-lingual, mid-lingual and disto-lingual) excluding third molars. We checked whether supragingival plaque (Pl) existed. The probing pocket depth (PPD) was determined by measuring the distance between the free gingival margin (FGM) and the base of the probing sulcus or pocket. Gingival recession (GR) was measured from the cementoenamel junction (CEJ) to the free gingival margin (FGM) and recorded as an integer: a positive value if the FGM was apical to the CEJ and a negative value if it was coronal to it. GR and PPD were added together to get the probing attachment level (PAL). BOP was marked as positive if bleeding started within 10–15 seconds after probing. Each patient’s BOP% (percentage of sites with BOP) and Pl% (percentage of sites with Pl) were calculated. The number of missing teeth (except for third molars), presence and absence of furcation and mobility were also recorded [17]. Results from duplicate examinations on 10 participants (every 4 from the third participant examined) showed that intra-examiner reliability on periodontal status (PPD/PAL in mm) were substantial (weighted Kappa = 0.618/0.810). Participants with any dental emergencies were immediately referred for proper management, while those with non-urgent oral problems were given referrals via the regular channels within the dental hospital.

### Sample collection

Subjects who qualified for the study were asked not to brush their teeth in the same morning when saliva/supra- or subgingival samples were taken which was within 1-2 weeks after the recruitment, screening and examination. Participants were asked not to eat nor drink (except water) for at least two hours before sample collection. Firstly, 2 ml of unstimulated saliva were collected according to Dawes 1972 [18]. For plaque collection, cotton rolls were used to isolate all teeth, which were air dried, and then pooled supragingival plaque were collected from intact tooth crowns (excluding any decayed and restored tooth-surfaces) using a sterile universal curette, followed by subgingival plaque sampling (pooled) using Gracey curettes from one deepest site on each quadrant.

In control participants, subgingival plaque was normally collected from a site with PPD of 3/4 mm, and supragingival plaque was collected from a buccal site at canine/premolar in each quadrant. In periodontitis participants, one deepest subgingival site per quadrant i.e. PPD ≥ 5 mm was sampled. In case the person had a quadrant without PPD ≥ 5 mm, an alternative non-adjacent deep pocket site (PPD ≥ 5 mm) in another quadrant was sampled as replacements. Immediately after sample collection, all specimens were transferred to sterile 2-ml cryovials and quickly frozen for long-term preservation at -80 C.

### DNA extraction

Frozen samples thawed on ice. Using the QIAamp DNA Mini kit (Qiagen), DNA was extracted according to the manufacturer’s instructions for gram-positive bacteria. 48 samples from three sites of the 39 subjects (16 healthy and 23 periodontitis) with purified DNA concentrations more than 50 ng/µL have been sent for further shotgun sequencing.

### NGS sequencing

Samples yielded sufficient amounts of DNA (≥ 50 ng/µL) have been selected to perform downstream shotgun metagenome sequencing. Library preparation (Pair-End sequencing of 151 bp) was performed at the Centre for PanorOmic Sciences (CPOS), Genomics Core (The University of Hong Kong, LKS Faculty of Medicine) using the Illumina NovaSeq 6000 platform. Libraries were prepared based on the KAPA HyperPrep Kit (KR0961) protocol. Fifty nanograms of genomic DNA was fragmented using the Diagenode Bioruptor Pico system to a peak size of around 300 bp.

End-repair, A-tailing at the 3’ end, and adaptor ligation with an integrated DNA technologies (IDT) dual-indexed unique molecular identifier (UMI) adaptor system was performed on the fragmented DNAs. Dual-Solid Phase Reversible Immobilization SPRI was used to select an adapter-ligated library ranging in size from 300 to 750 bp. PCR was performed to enrich the libraries. For quality control analysis, Agilent Bioanalyzer, Qubit, and qPCR were used to validate the enriched libraries. The libraries were denatured, and their concentration was optimized.

### BGC identification of oral bacteria

After generating the raw short reads, the KneadData pipeline was used to trim the barcodes, filter out low-quality reads and remove human reads from the samples. We used trimmomatic settings for a 4-base wide sliding window, with a minimum length of 90 bp and an average quality per base greater than 20. The final high-quality reads were assembled using MEGAHIT to generate the long contigs [19]. antiSMAH 6.0 was then used to identify BGCs within the long contigs [20]. antiSMASH detects BGCs consisting of multiple gene clusters of different types merged into a single large gene cluster. We developed a code to convert GenBank to FASTA using BioPython [21] python library to facilitate further bioinformatic analysis. Based on antiSMASH glossary [20] we mapped the discovered BGCs into 6 super-classes, namely, non-ribosomal peptides (NRPs), polyketides, ribosomally encoded and post-translationally modified peptides (RiPPs), saccharides, terpenes, and others (Table S1).

### Taxonomic assignment and phylogenetic tree

Taxonomic assignment of the assembly contigs were estimated by CAT (setting -sensitive -r 10 and -f 0.3) [22] based on the Genome Taxonomy Database (GTDB) [23]. Phylogenetic trees were constructed using PhyloT (https://phylot.biobyte.de) which were visualized using iTol (https://itol.embl.de/).

### BGCs normalized abundance calculation

Bowtie2 [24] was used to quantify the number of hits to each BGC. We parsed the SAM output file from Bowtie2 to extract number of hits. We considered hits with an alignment length more than 60 bp and sequence identity more than 0.95. We calculated the normalized abundance of each BGC *I* in each sample *j* using the following equation:

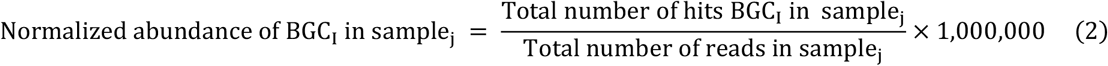

### BGCs novelty score calculation

To calculate BGC novelty, Biosynthetic Genes Super-Linear Clustering Engine (BiG-SLiCE) was executed in -query mode using a previously prepared dataset consisting of 1.2 million BGCs [25]. The resulting distance *d* represents the degree of similarity between a particular BGC and previously computed gene cluster families (GCFs), with a larger *d* suggesting greater uniqueness. For this research, we classified BGCs with a BiG-SLiCE score (*d*) greater than 1500 as novel BGCs. We used blast2GO [26] to perform gene ontology analysis of the novel unique clusters.

### BGCs site-specificity analysis

To facilitate comparisons, all sequences obtained for a specific domain were pooled and clustered according to sequence identity. We clustered the BGCs using CD-Hit (similarity > 0.5) [27]. We classified the representatives BGCs based on whether or not they are in saliva, subgingival, supragingival, or all three sites. For each oral site, the frequency of each of the representatives BGCs was calculated. We used Venn diagram to plot the result.

### BGC network analysis

To quantify the diversity of the BGCs, we analysed all BGCs with BiG-SCAPE [28], which groups similar clusters into a gene cluster family (GCF). BiG-SCAPE generates a network based on the sequence similarity between the discovered BGCs and 1,409 experimentally validated BGCs from the MIBIG repository [29]. We used domain sequence similarity score (weighted by sequence identity), to define a final distance metric for pairwise comparisons between BGCs. We saved the network in CSV format and visualized with Cytoscape [30].

**<numbers are not consistent!!>**

## Results

We collected oral microbial samples from 3 distinct oral niches: 1) unstimulated saliva, 2) supragingival plaque (multi-site, full-mouth), and 3) sub-gingival plaque (the deepest site per quadrant, pooled) from 39 subjects who were suffering from periodontitis (n = 23), or were controls without severe periodontitis nor advanced periodontal attachment loss (n = 16) [31]. All participants were ethnic Chinese living in Hong Kong. 48 samples with sufficient amounts of DNA (≥ 50 ng/µL) were selected for downstream metagenome sequence analysis. Details of the samples sequenced are summarized in (**Tables 1 and Table S2)**.

**Table 1.** Summary of 48 clinical samples analyzed by shotgun metagenomic sequencing.

|  | Saliva | Subgingival<br>plaque | Supragingival<br>plaque | Total |
| --- | --- | --- | --- | --- |
| Control subjects | 11 | 3 | 4 | 18 |
| Periodontitis subjects | 9 | 11 | 10 | 30 |
| Total | 20 | 14 | 14 | 48 |

### BGC profile across the oral bacterial metagenome

Shotgun sequencing of the 48 selected samples produced an average of 94,076,902 ± 6,030,847 reads per sample, ranging from 79,059,540 to 108,878,790 (**Table S3**). We removed the human reads from each sample and assembled the remaining short reads to construct bacterial genome contigs. Then, we analyzed the bacterial genome contigs using the antiSMASH 6.0 program to identify the BGCs present [20]. We discovered 10,742 BGCs from 44 distinct groups (**Table S4**). These BGCs were divided into six ‘super-classes’: non-ribosomal peptides (NRPs), polyketides (PKs), ribosomally-encoded and post-translationally modified peptides (RiPPs), saccharides, terpenes (isoprenoids), and “Others” based on antiSMASH glossary [20] (**Figure 1A**). A significant proportion (3,833 or 36% of total BGC identified) of the identified BGCs were labelled as polyketides, making it the most prevalent type of BGC in the oral cavity. Polyketide pathways were found in taxa assigned to diverse bacterial phyla, but were most common in Bacteroidetes, Proteobacteria, Actinobacteria, Firmicutes, and Fusobacteria; with Bacteroidetes taxa having the most (**Figure 1B**). Aryl polyenes (APEs) were the most prevalent polyketide subclass (2,983, 77% of total polyketide identified).

**Figure 1.**
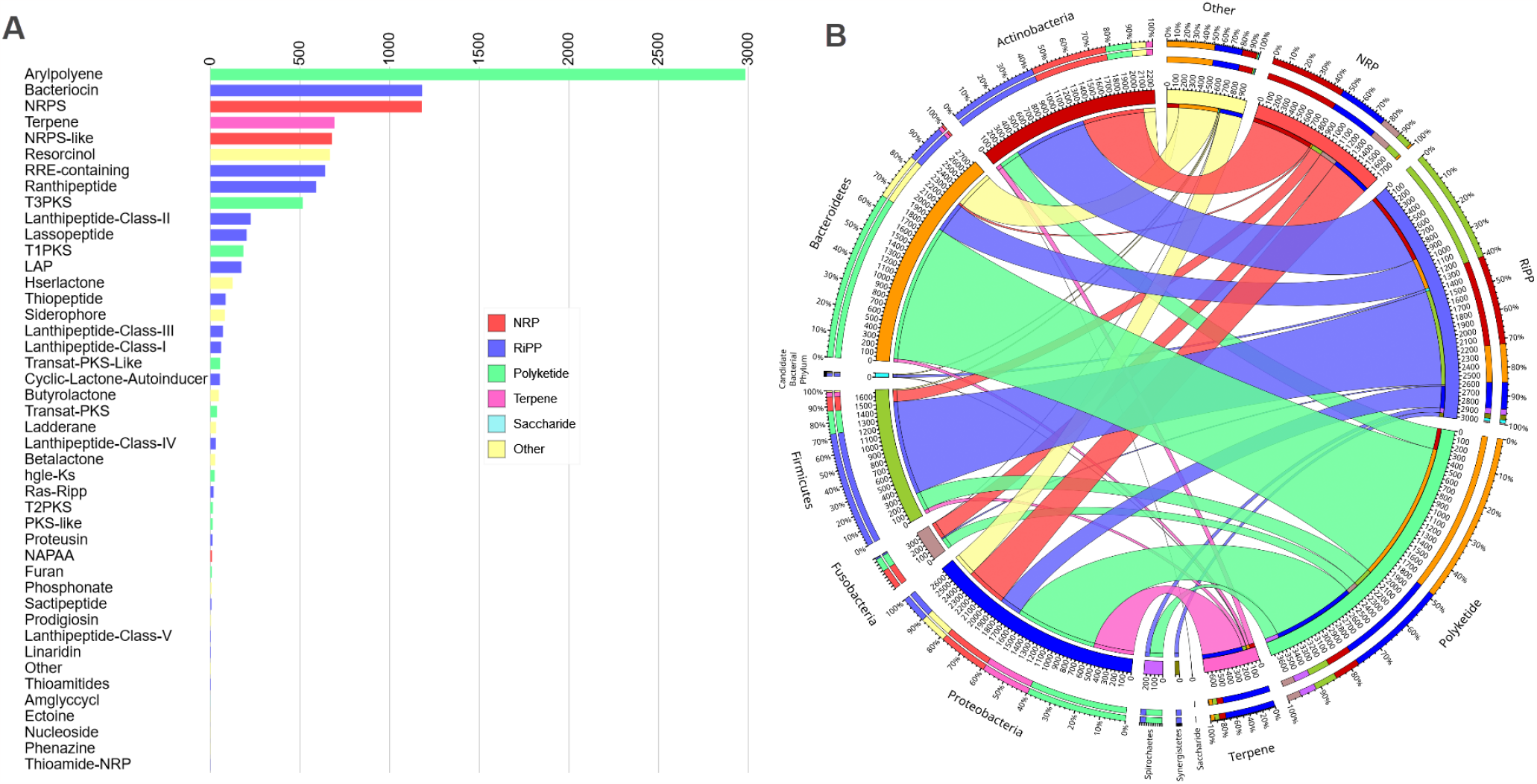
BGCs profile in the 48 oral samples. (A) 44 different sub-types of BGCs and their frequency in the oral samples collected. (B) The relationship between BGC super-class and their bacterial phylum sources.

Ribosomally-synthesized and post-translationally modified peptides (RiPPs) were another frequently-detected BGC super-class, corresponding to 3,355 BGCs of a known class (31% of total BGC identified). We identified several different ‘sub-types’ of RiPP BGCs in clinical samples including bacteriocins, lantipeptides, ranthipeptides, sactipeptides, and proteusins (**Figure 1A**). Bacteriocin was the most prevalent BGC among the RiPP sub-types, accounting for 1,181 BGCs (35% of all identified RiPP). RiPP-producing BGCs were widespread across diverse bacterial taxa, with most identified within Firmicutes genomes (**Figure 1B**). In addition, we found a variety of BGCs encoding non-ribosomal peptide synthetases (NRPS; 1,866 BGCs; 17% of all BGCs identified) in the oral cavity. We also found 692 (7% of all BGCs identified) Terpene-class BGCs in our samples. The rare classes of BGCs which were categorized as “Others” comprised 995 BGCs (9% of all BGCs identified).

When the BGC counts were broken down by BGC type and phylum, it was discovered that the three well-known producer phyla, *Bacteroidota (Bacteroides), Proteobacteria*, and *Actinomycetota (Actinobacteria)*, contributed more than 70% of the BGCs. *Firmicutes* and *Fusobacteria* BGCs accounted for *ca*. 20% of total BGCs, other phyla accounted for the remaining 4%; with 6% of BGCs remained unclassified at the phylum level (**Figure S1 A, B)**.

### BGC abundance in control and periodontitis subjects

We next looked at how the bacterial BGCs differed between control and periodontitis patients. We calculated the normalized abundance of each BGC in each sample (see Methods). The normalized abundance of each BGC in the case and control groups were compared using Wilcoxon–Mann– Whitney test. BGCs with different abundances in control versus periodontitis were linked to the synthesis of a wide range of small molecule types, from different bacterial taxa (**Figure S2**). We ranked the BGCs based on the p-value and the heat-map plot of the top twenty BGCs enriched in control and periodontitis samples showed in **Figure 2A**. For example, BGCs encoding resorcinol, thiopeptide, and NPRs from *Porphyromonas gingivalis, Treponema denticola* and *Desulfobulbus oralis* were more frequently detected in patients with periodontitis, while BGCs encoding NRPS, T3PKS and betalactone from *Rothia mucilaginosa, Streptococcus salivarius* and *Haemophilus parainfluenzae* were more frequently detected in samples from control participants (Figure 2 B-G).

**Figure 2.**
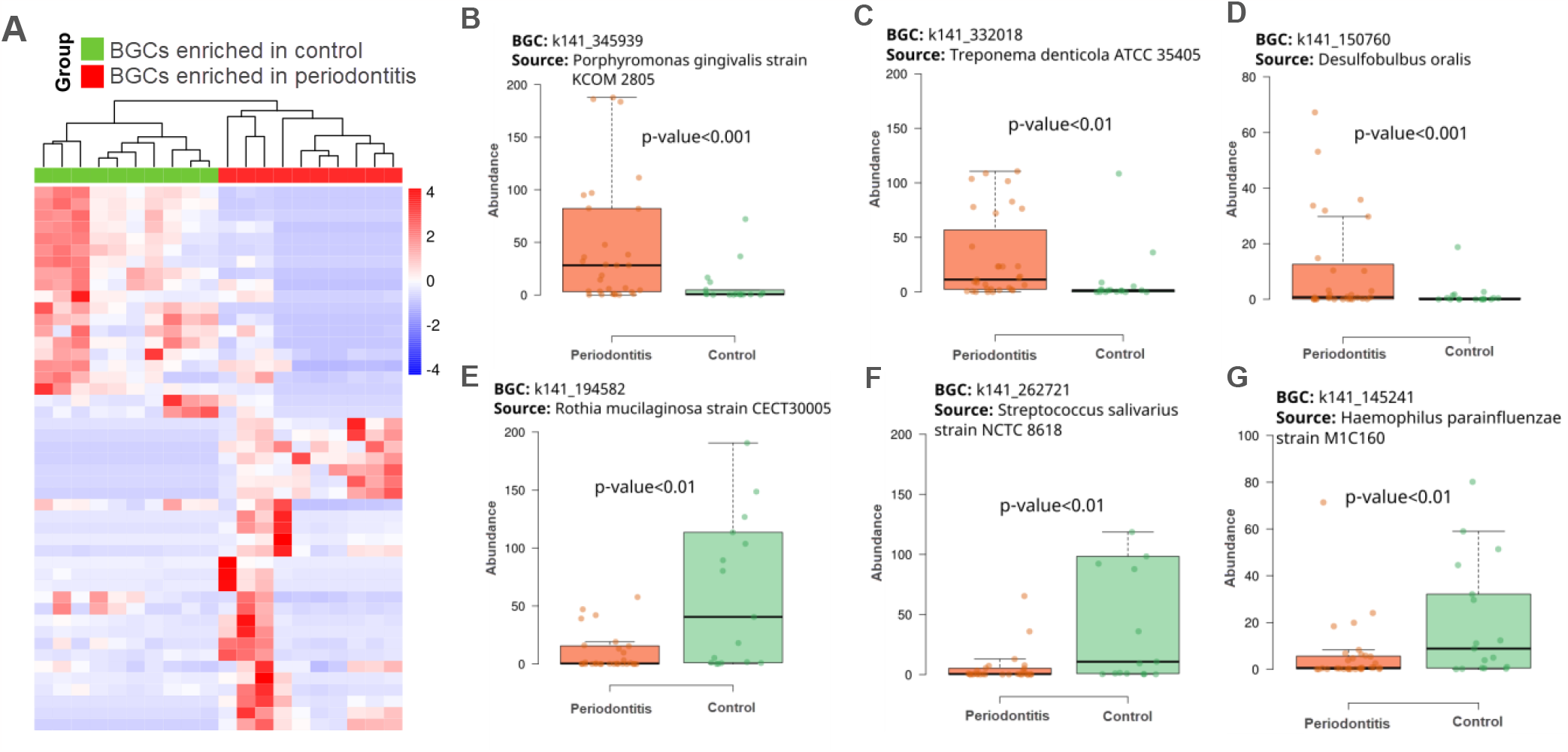
Comparison of the BGCs between control and periodontitis oral samples. (A Heat-map showing the differences in distributions of the top twenty BGCs between control and periodontitis subjects. (B-D) Identities of 3 BGCs that were enriched in the periodontitis cases. (E-F). Identities of 3 BGCs that were enriched in the control cases.

### The novelty of the oral BGCs

We used BiG-SLiCE in query mode to calculate the distance (*d*) of discovered BGCs sequences with a set of 1.2 million known BGCs [25]. There was a wide range of distances (*d*) within each BGCs class, indicating that each class included BGCs that were both closely and distantly connected to previously identified BGCs (**Figure 3A**). Our results showed that 207 BGCs had a *d* > 1500, which showed they were extremely divergent from known BGCs (**Table S5**). The Polyketide BGC cluster had a higher *d* score, with 125 novel BGCs (score > 1500). After that, the NRP and Terpene BGCs respectively contained 41 and 23 novel clusters. Among the novel BGCs, 61% of them appeared in the periodontitis samples, and 39% in the control samples (**Figure 3B**). Moreover, Proteobacteria, Bacteroidetes and Actinobacteria were the top three bacterial phylum containing these novel BGCs (**Figure 3C**).

**Figure 3.**
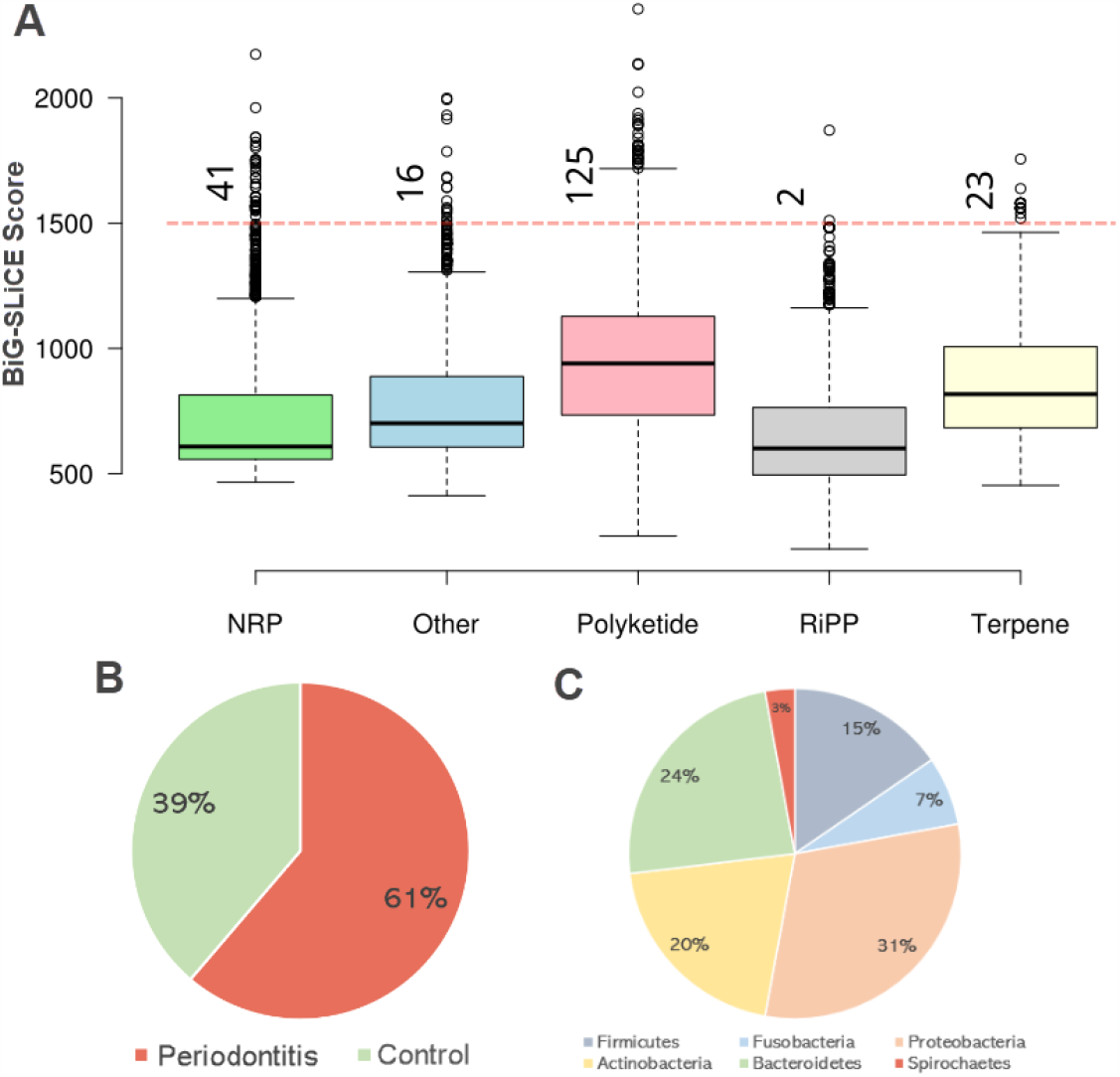
Novelty of oral BGCs identified in periodontitis and control subjects. (A) boxplot of BiG-SLiCE scores of BGCs. The dash red line shows the threshold (d>1500). (B) Proportions of the 207 novel BGCs within control and periodontitis samples (C) The bacterial sources (genomic hosts) of the 207 novel BGCs

### Oral BGC site specificity

We investigated if the discovered BGCs were site-specific by clustering them based on the site of detection. For comparison, all the BGCs sequences were pooled and then clustered based on sequence similarity. The resulting clusters were classified based on whether they contained sequences from saliva, subgingival plaque, supragingival plaque, or all three sites (see Methods). We then used Venn diagrams to display the extent of overlap between different oral sites (**Figure 4A**).

**Figure 4.**
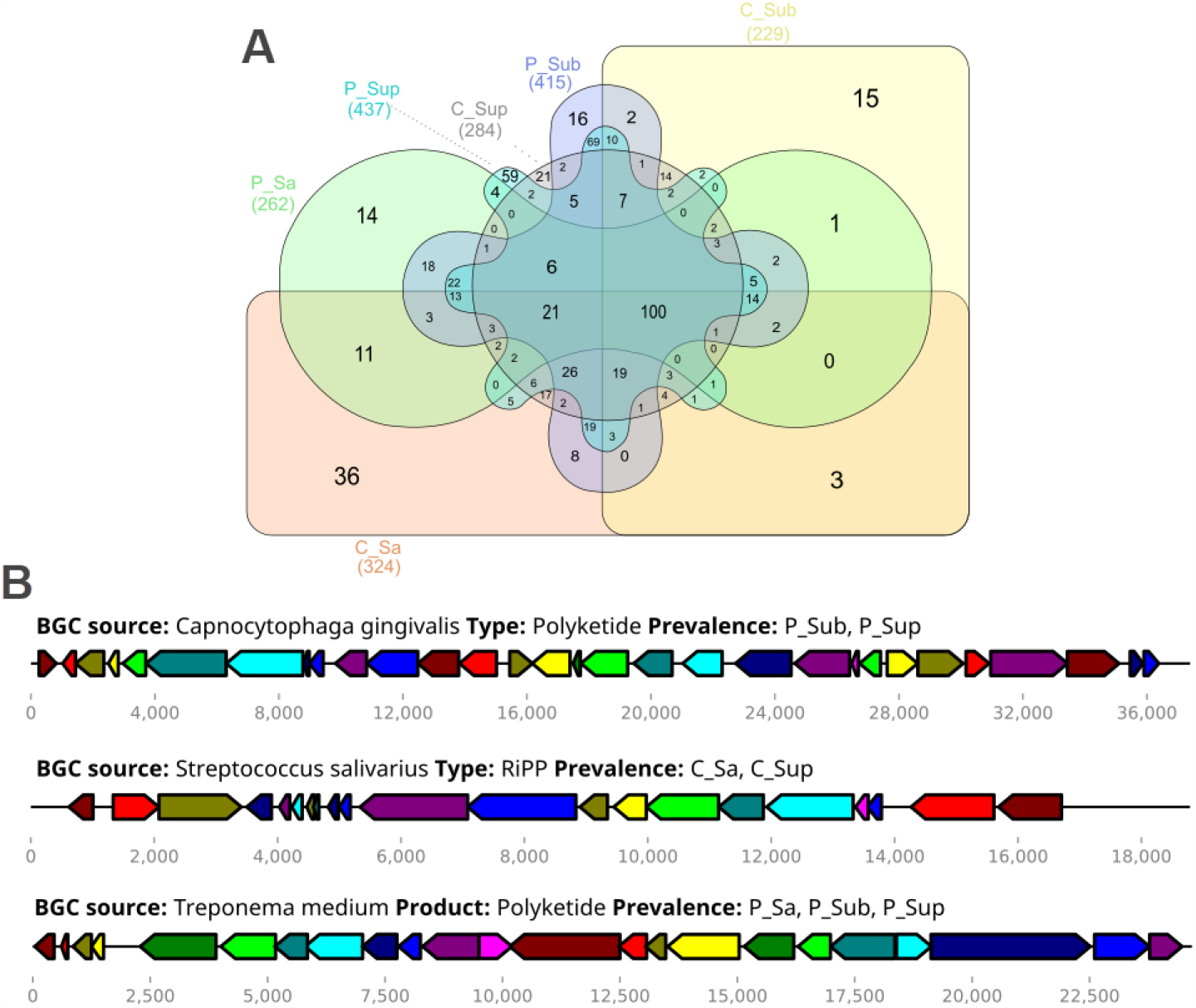
Site-specificity of oral BGCs. (A) Venn diagrams representing BGCs present in the six different groups. (C_Sa: control saliva sample, C_Sup: control supragingival sample, C_Sub: control subgingival sample, P_Sa: periodontitis saliva sample, P_Sub: periodontitis subgingival sample, P_Sup: periodontitis supragingival sample); (B) Three selected BGCs from the oral microbiota, spanning in samples from different oral niches.

For example, we found that a BGC putatively encoding a Polyketide metabolite (BGC cluster id: k141_164411) present within *Capnocytophaga gingivalis* was only present in the supragingival and subgingival plaque of periodontitis subjects (n = 7), whilst a BGC putatively encoding a RiPP (BGC cluster id: k141_302416) present within *Streptococcus salivarius* contigs, was only present in saliva from controls. In contrast, some BGCs were prevalent in all three oral sites, such as the BGC encoded by *Treponema medium* (BGC cluster id: k141_23213), which was found in all three sites of periodontitis patients’ oral sites (n = 15) (**Figure 4B**).

### Network analysis of oral BGCs

We performed network analysis to identify potentially clinically-important bacterial BGCs. We measured the pairwise distance of BGCs found in our samples from known gene cluster families (GCFs) to categorize the BGCs into families. We converted the BGCs to arrays of protein domains, then compared them based on the proportion of shared domains and the sequence similarities. We clustered both whole and fragmented BGCs into gene cluster families using BiG-SCAPE and CORASON [28] to gain a comprehensive overview of BGC diversity in large genome collections.

The resulting network contained 5,267 nodes and 22,643 connecting edges for periodontitis and 2,893 nodes and 8,728 connecting edges for control samples, indicating both close and distant homology to known biosynthetic pathways (**Figures S3, S4**). Polyketide BGCs made up the majority of the largest subnetwork, whereas RiPP BGCs were the primary constituents of the second-largest subnetwork. In order to make it easier to visualize these large networks, we determined the hub nodes which had the highest connectivity within the network. For example, BGC: K141_4883 was a hub BGC in the control network and K141_1504 and K141_316297 were hub BGCs in the periodontitis network. We used ClusterBlast to compare these hub BGCs with the known BGCs in the MiBIG databases [29]. K141_4883 had a 61% similarity with a previously identified BGC from *Streptococcus salivarius*, K141_1504 had a %52 similarity with a BGC from *Centipeda periodontii and* K141_316297 had a 52% alignment similarity with a BGC from *Porphyromonas gingivalis* (**Figure 5**).

**Figure 5.**
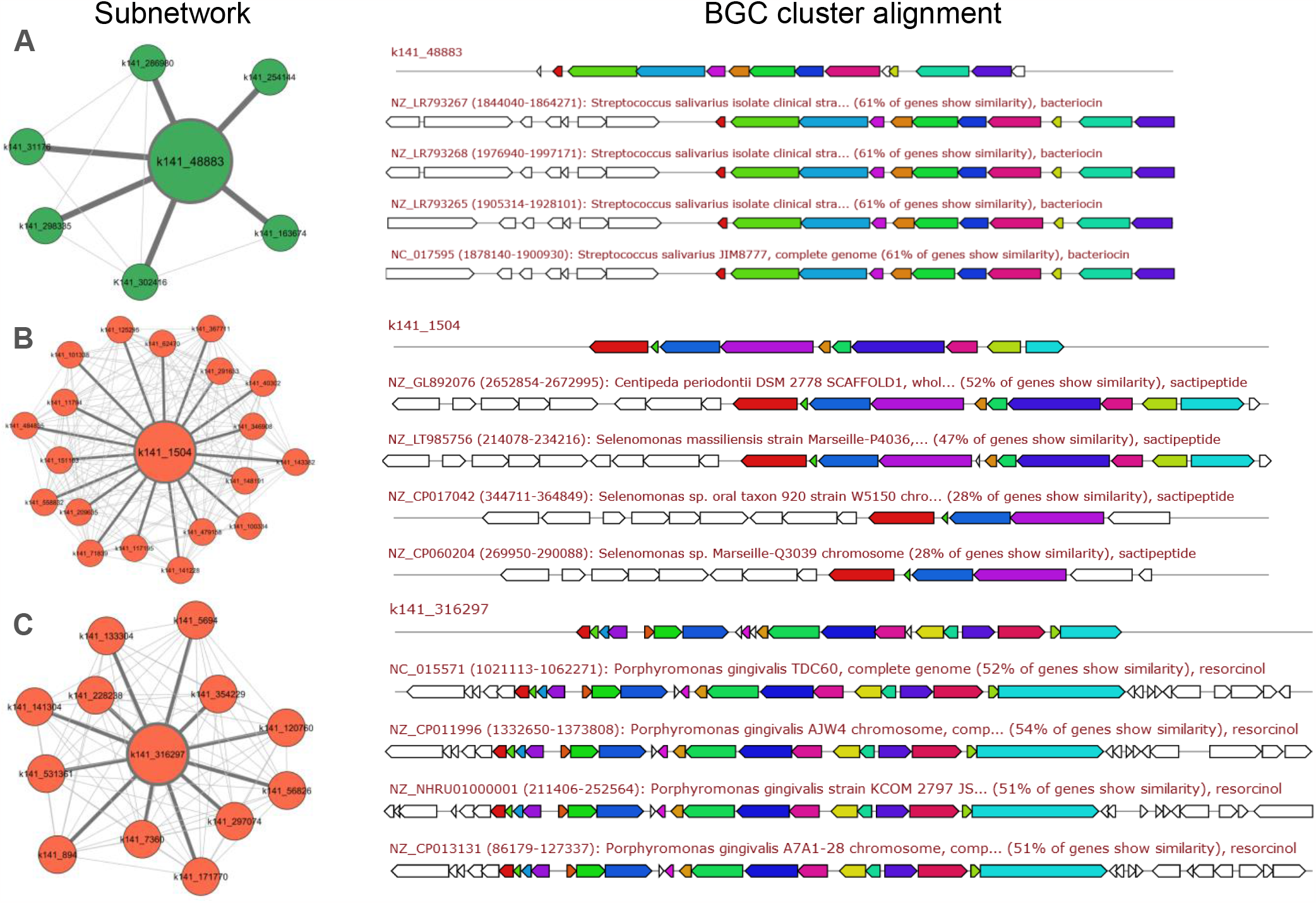
Network analysis of selected BGCs. The hub BGC nodes in control and periodontitis networks.

Comparison of these hub nodes with BGCs with known secondary metabolite products enabled us to identify their putative products. Our comparison showed that the putative product of K141_4883 was similar to the salivaricin CRL1328 α-peptide (score: 0.52). K141_1504 was predicted to produce a compound similar to huazacin (score: 0.42) and the putative product of K141_316297 was the molecule SRO15-3108 (score: 0.53) (**Table S6**).

## Discussion

Our results show that the oral cavity contains many different types of BGCs encoded by diverse bacterial taxa. APEs are polyunsaturated carboxylic acids that were discovered to be the result of the most common family of bacterial BGCs in our samples. APEs can be found in a variety of gram-negative host-associated bacteria, including multidrug-resistant human infections. Many anaerobes populate the gingiva and tongue, which have a high concentration of APEs BGCs [32]. Studies reveal that APEs in outer membrane of the bacteria induces biofilm formation by modification of regulatory cascades or cell envelope composition [33].

BGCs predicted to synthesize bacteriocins were the second most abundant type of BGCs in our cohort. Bacteriocins are produced by Gram-positive and Gram-negative bacterial species, and may exhibit narrow or broad spectrum antimicrobial activities [34, 35]. They exhibit antagonistic behaviours against a range of bacterial taxa, and affect competition for persistence between commensals and pathogens, thus having the ability to elicit large-scale changes in microbiome structure [9, 36]. Bacteriocin BGCs are typically highly transmissible (via horizontal gene transfer) and may evolve rapidly [35]. Oral bacteria (especially streptococci and lactobacilli) are known to produce and secrete a range of bacteriocins, which may help reduce populations of cariogenic bacteria and respiratory pathogens [37, 38]. Bacteriocin, on the other hand, is thought to be an important factor in the colonization and establishment of *Streptococcus mutans* in the dental biofilm [39] which is associated to chronic periodontitis and periodontal parameters [40].

We found a higher abundance of BGC from *P. gingivalis* in periodontitis samples. *P. gingivalis* is a species of bacteria identified to be linked to periodontitis [41]. This BGC was predicted to synthesize resorcinol which is a phenolic compound. Studies showed a syntrophic interaction between resorcinol-degrading bacteria (RDB) and anode-respiring bacteria (ARB) was involved in biofilm formation [42]. Moreover, BGCs from *T. denticola* and *D. oralis*, which are predicted to produce thiopeptide and NRP-like compounds, respectively, were observed to be more abundant in periodontitis samples. Thiopeptides may function as antibiotic agents, however their roles in a complex microbial community remains unclear. Studies have shown that thiopeptides may have an influence on neighbouring microorganisms in a microbial niche to stimulate biofilm development [43]. In contrast, expression of BGCs encoding NRPS, T3PKS, and betalactone from *Rothia mucilaginosa, Streptococcus salivarius*, and *Haemophilus parainfluenzae* were higher in control samples. Studies show that *Rothia mucilaginosa*, a common resident of the oral cavity, is an anti-inflammatory bacterium which has an inhibitory effect on pathogen-or lipopolysaccharide-induced pro-inflammatory responses [44]. Likewise, studies show an anti-inflammatory properties for *Streptococcus salivarius* [45]. In addition, *Haemophilus parainfluenzae* was found in greater abundance in control samples than in periodontitis samples in studies [46].

The comparison of BGCs we discovered with known clusters helped us in identifying unique clusters not represented in previous studies. We found that polyketide and NRP are two classes of BGCs with the highest number of novel clusters in our oral cavity samples. Around 60% of these unique novel BGCs belong to periodontitis patients. The gene ontology analysis of these unique novel cluster shows that they play a key role in acyltransferase activity and metal ion binding activity (Figure S5). To the best of our knowledge, this is the first study to find BGCs with possibly unique features in the oral microbiota. These novel BGCs can be explored further for their roles in periodontitis, which could lead to the discovery of new therapeutic to promote oral health. Besides, our findings suggest that BGCs may be selectively present in specific oral sites (niches). This finding is aligned with the concept that microbial communities of the mouth may be highly site- (niche)-selective [47, 48]. Different oral sites (e.g. specific tooth surfaces, individual healthy or inflamed gingival sulci) may provide distinct biological environments that strongly promotes the growth of certain bacterial consortia over others [10].

However, there are three significant limitations to our study. First, our analysis is limited by the small sample sizes for each oral site. This was largely due to the low quantities of DNA we could extract from many clinical samples, which precluded their metagenomic analysis. Second, it is important to highlight that the estimated BGC diversity indicated here is likely underestimated. This is due to the fact that the resolution of shotgun metagenome sequencing may not be appropriate to capture all strain-level BGCs. Third, our data and analysis are limited to the bacterial genome level. To better understand the role of BGCs and their products, metatranscriptomic analysis is required.

## Conclusion

Our findings emphasize the presence of hundreds of widely distributed BGCs with unknown functions in the human oral microbiome. These BGCs can be used as a resource for future research that is looking to identify small-molecule-mediated interactions in the human oral microbiota.

## Data Availability

All data produced are available online at NCBI database (BioProject accession numbers: PRJNA932553).

https://www.ncbi.nlm.nih.gov/bioproject/PRJNA932553

https://figshare.com/projects/Systematic_Analysis_of_Site-Specific_Biosynthetic_Gene_Clusters_BGCs_of_Oral_Microbiome_in_Periodontitis/157008

## Data availability

All shotgun metagenome data described in this study were submitted to the Short Reads Archive database (BioProject accession numbers: PRJNA932553). The sequence of all BGCs and the result of antiSMASH output can be found in https://figshare.com/projects/Systematic_Analysis_of_Site-Specific_Biosynthetic_Gene_Clusters_BGCs_of_Oral_Microbiome_in_Periodontitis/157008.

## Author Contributions

MK, WKL and RMW designed the study; WKL collected samples; MK analyzed data; MK, RMW and WKL wrote the paper; MK contributed to funding.

