## Supplementary information for "Multi-Site Analysis of Biosynthetic Gene Clusters (BGCs) from the Periodontitis Oral Microbiome"

**Supplementory Figures:**

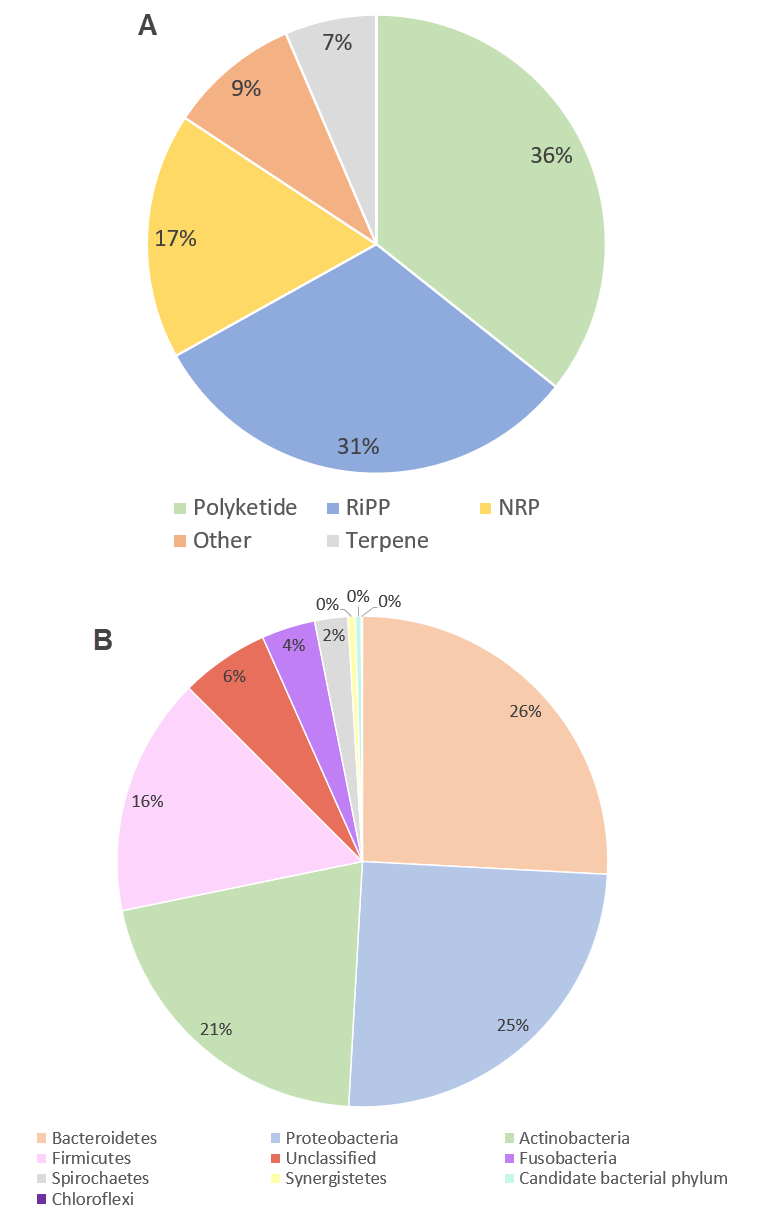

**Figure S1. Oral biosynthetic gene clusters (BGCs) profile of samples with purified DNA concentration ≥ 50 ng/µL.** (A) distributation of the BGCs based on their product type (B) distributation of the BGCs based on their bacterial phylum

| 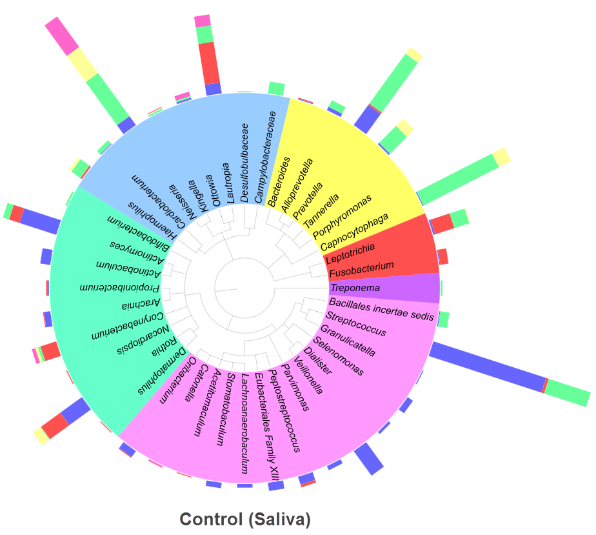 | 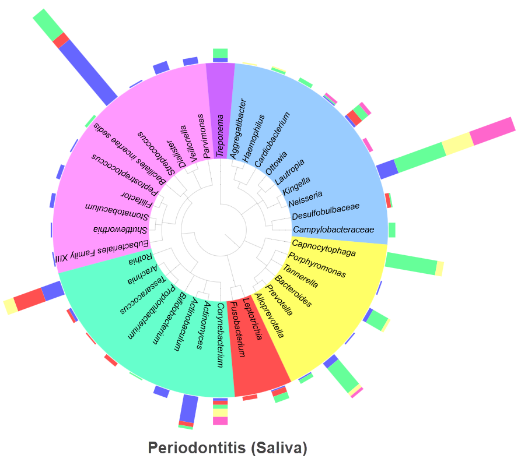 |
| --- | --- |
| 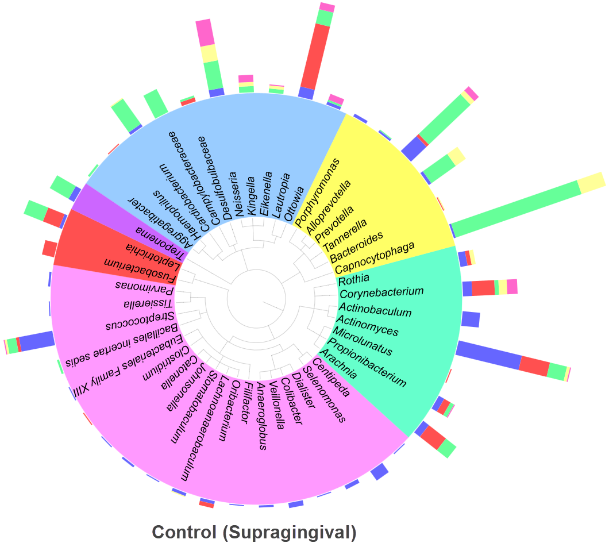 | 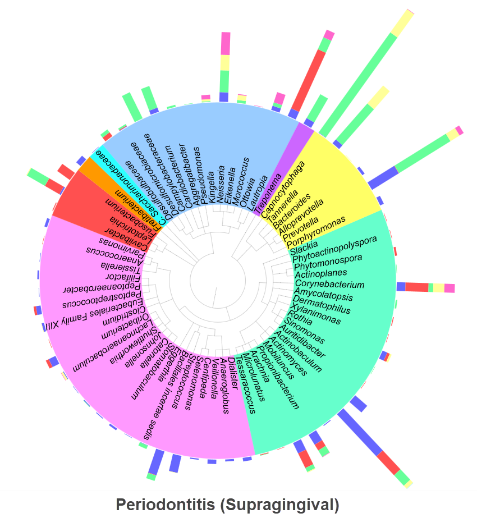 |
| 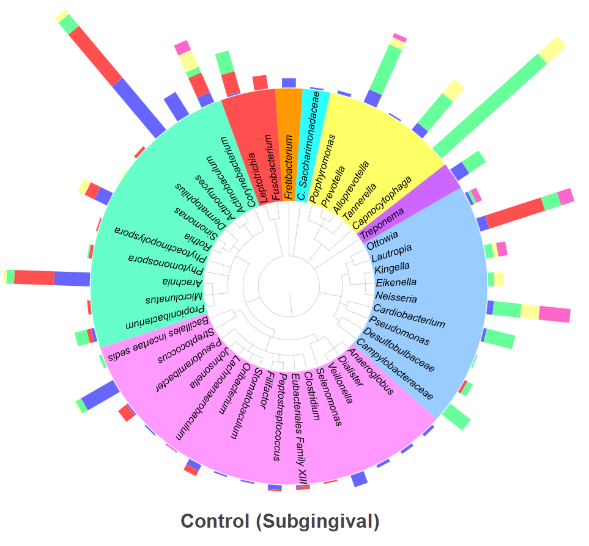 | 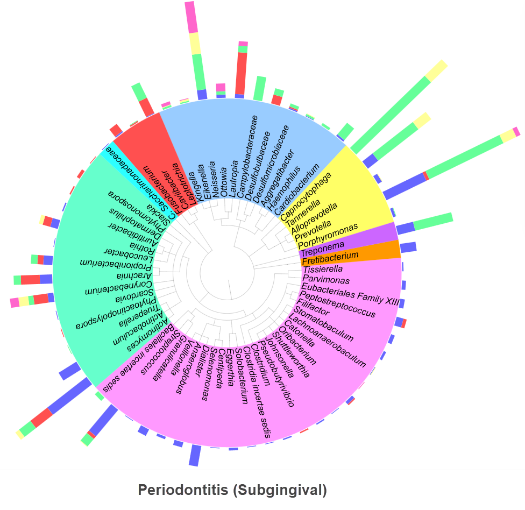 |
| 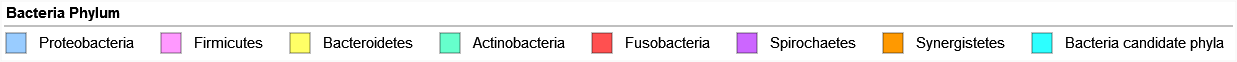 | |
| 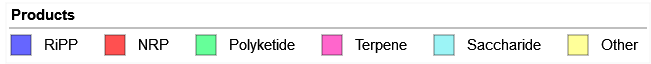 | |
| **Figure S2.** Phylogenetic tree of the bacterial genus tentatively produced the biosynthetic gene clusters (BGCs) in control and periodontitis samples with purified DNA concentration ≥ 50 ng/µL. | |

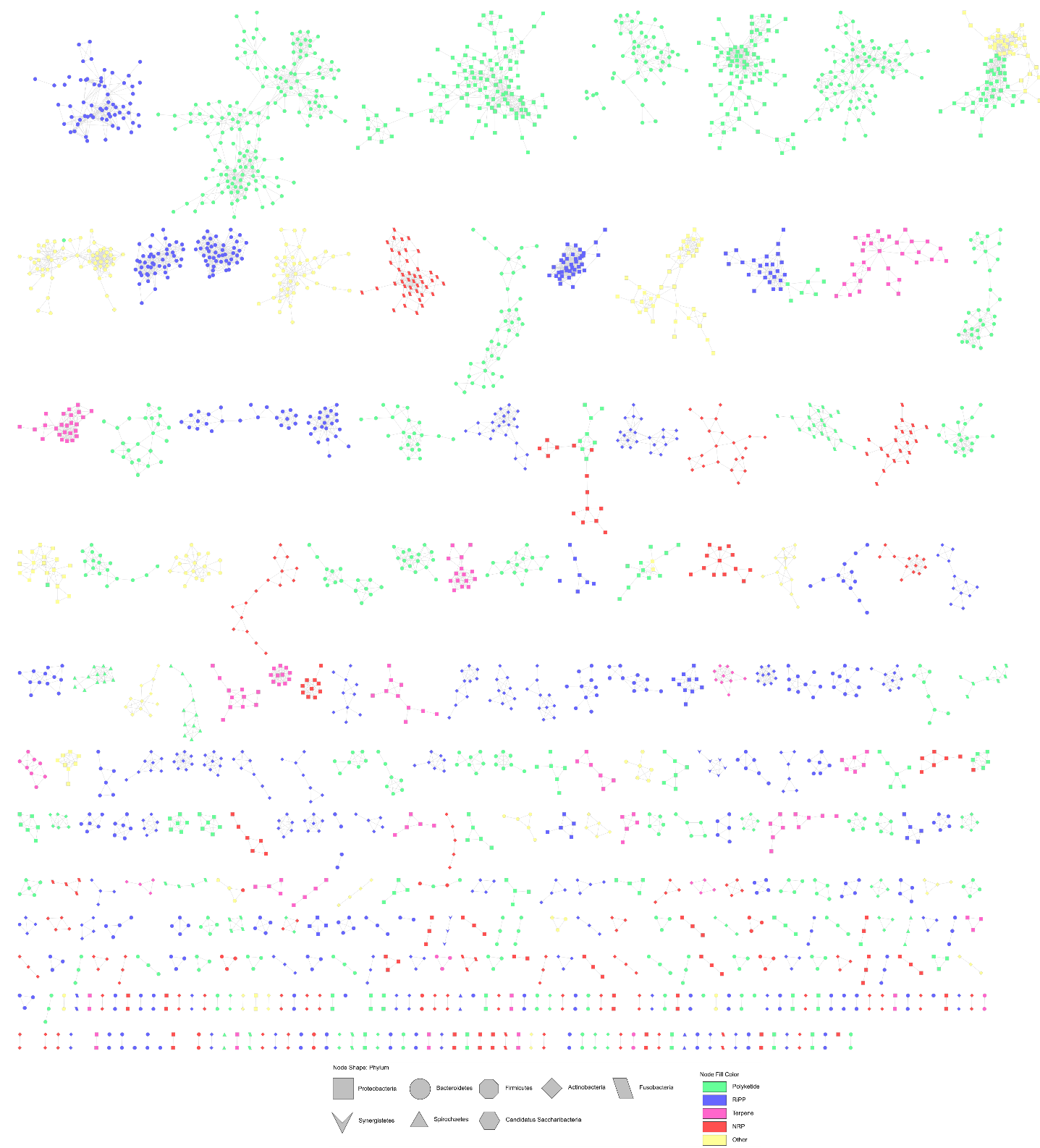

**Figure S3. Similarity networks of the biosynthetic gene clusters (BGCs) identified in control participants.** Edges drawn between the nodes correspond to pairwise distances, computed by Biosynthetic Gene Similarity Clustering and Prospecting Engine (BiG-SCAPE) as the weighted combination of the Jaccard, adjacency, and domain sequence similarity indices.

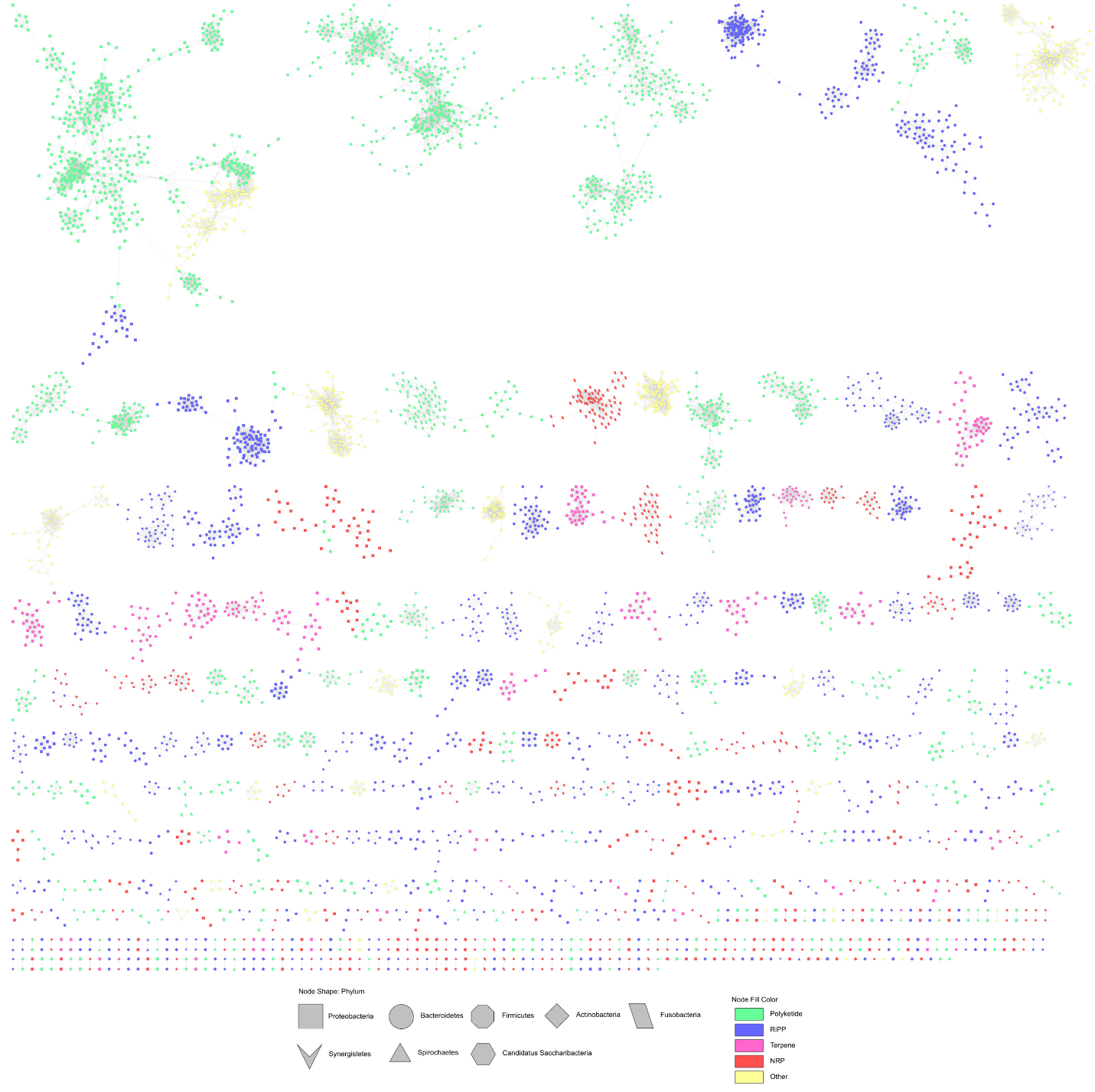

**Figure S4. Similarity networks of the biosynthetic gene clusters (BGCs) identified in oral samples with periodontitis.** Edges drawn between the nodes correspond to pairwise distances, computed by Biosynthetic Gene Similarity Clustering and Prospecting Engine (BiG-SCAPE) as the weighted combination of the Jaccard, adjacency, and domain sequence similarity indices.

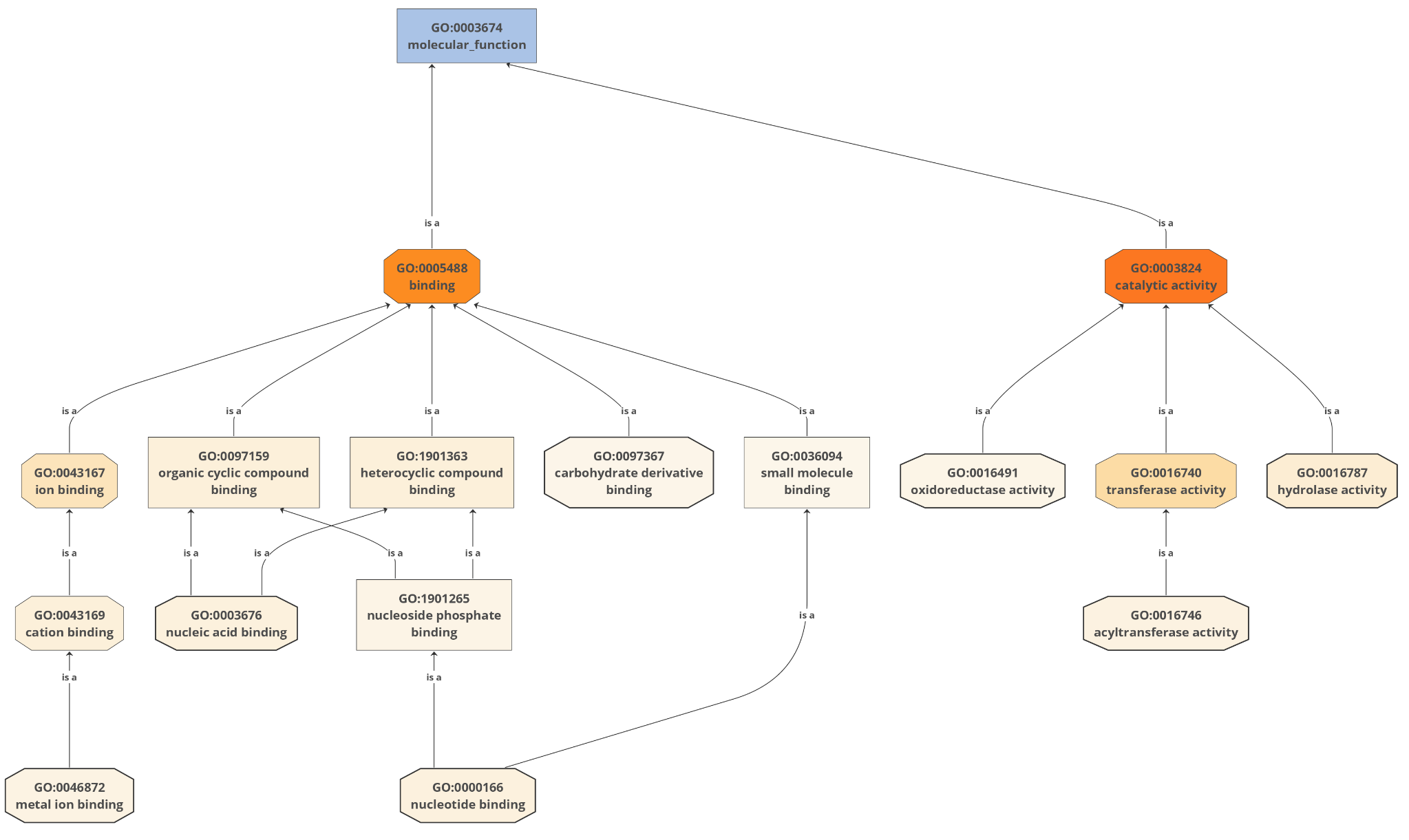

**Figure S5.** Gene ontology analysis of the unique novel BGCs.

**Supplementory Tables:**

**Table S1.** The 44 biosynthetic gene clusters (BGCs) subtypes identified and their mapped superclass (map to six super classes)

| **BGCs** | **Super class** |
| --- | --- |
| Arylpolyene | Polyketide |
| Ripp-Like | RiPP |
| NRPS | NRP |
| Terpene | Terpene |
| NRPS-like | NRP |
| Resorcinol | Other |
| RRE-containing | RiPP |
| Ranthipeptide | RiPP |
| T3PKS | Polyketide |
| Lanthipeptide-Class-II | RiPP |
| Lassopeptide | RiPP |
| T1PKS | Polyketide |
| LAP | RiPP |
| Hserlactone | Other |
| Thiopeptide | RiPP |
| Siderophore | Other |
| Lanthipeptide-Class-III | RiPP |
| Lanthipeptide-Class-I | RiPP |
| Transat-PKS-Like | Polyketide |
| Cyclic-Lactone-Autoinducer | RiPP |
| Butyrolactone | Other |
| Transat-PKS | Polyketide |
| Ladderane | Other |
| Lanthipeptide-Class-IV | RiPP |
| Betalactone | Other |
| hgle-Ks | Polyketide |
| Ras-Ripp | RiPP |
| T2PKS | Polyketide |
| PKS-like | Polyketide |
| Proteusin | RiPP |
| NAPAA | NRP |
| Furan | Polyketide |
| Phosphonate | Other |
| Sactipeptide | RiPP |
| Prodigiosin | Other |
| Lanthipeptide-Class-V | RiPP |
| Linaridin | RiPP |
| Other | Other |
| Thioamitides | RiPP |
| Amglyccycl | Saccharide |
| Ectoine | Other |
| Nucleoside | Other |
| Phenazine | Other |
| Thioamide-NRP | RiPP |

**Table S2.** Periodontal disease diagnosis of study participants (n = 39) whose sample(s) were with purified DNA concentration ≥ 50 ng/µL (n = 48).

**Table S3.** Summary of the 48 shotgun metagenome samples from the 39 study participants (attached as xlsx file)

**Table S4.** List of the 10,742 bacterial biosynthetic gene clusters (BGCs) identified from the 48 oral samples with purified DNA concentration ≥ 50 ng/µL (attached as xlsx file)

**Table S5.** Novel oral biosynthetic gene clusters (BGCs): those with Biosynthetic Genes Super-Linear Clustering Engine (BiG-SLiCE) score > 1500 (n = 207) (attached as xlsx file)

**Table S6.** Comparison of the hub biosynthetic gene clusters (BGCs) with BGCs with known secondary metabolites from Minimum Information about a Biosynthetic Gene cluster (MiBIG)

| **Discovered BGC** | **Known BGC** | **Similarity score** | **Type** | **Compound(s)** | **Organism** |
| --- | --- | --- | --- | --- | --- |
| **K141_48883** | [BGC0000624.1](https://mibig.secondarymetabolites.org/repository/BGC0000624/index.html#r1c1) | 0.52 | RiPP | salivaricin CRL1328 α peptide, salivaricin CRL1328 β peptide | *Lactobacillus salivarius* |
| **k141_1504** | [BGC0001887.1](https://mibig.secondarymetabolites.org/repository/BGC0001887/index.html#r1c1) | 0.42 | RiPP | huazacin | *Bacillus thuringiensis* serovar *huazhongensis* BGSC 4BD1 |
| **k141_316297** | [BGC0000554.1](https://mibig.secondarymetabolites.org/repository/BGC0000554/index.html#r1c1) | 0.53 | RiPP | SRO15-3108 | *Streptomyces filamentosus* NRRL 15998 |
